# A CCG expansion in *ABCD3* causes oculopharyngodistal myopathy in individuals of European ancestry

**DOI:** 10.1101/2023.10.09.23296582

**Authors:** Andrea Cortese, Sarah J Beecroft, Stefano Facchini, Riccardo Curro, Macarena Cabrera-Serrano, Igor Stevanovski, Sanjog Chintalaphani, Hasindu Gamaarachchi, Ben Weisburd, Chiara Folland, Gavin Monahan, Carolin K Scriba, Lein Dofash, Mridul Johari, Bianca R Grosz, Melina Ellis, Liam G Fearnley, Rick Tankard, Justin Read, Melanie Bahlo, Ash Merve, Natalia Dominik, Elisa Vegezzi, Ricardo P Schnekenberg, Gorka Fernandez, Marion Masingue, Diane Giovannini, Martin Delatycki, Elsdon Storey, Mac Gardner, David Amor, Garth Nicholson, Steve Vucic, Robert D Henderson, Thomas Robertson, Jason Dyke, Vicki Fabian, Frank Mastaglia, Mark R Davis, Marina Kennerson, OPDM study group, Genomics England, Ros Quinlivan, Simon Hammans, Arianna Tucci, Catriona A McLean, Nigel G Laing, Tanya Stojkovic, Henry Houlden, Michael G Hanna, Ira Deveson, Paul J Lockhart, Phillipa J Lamont, Michael C Fahey, Enrico Bugiardini, Gianina Ravenscroft

## Abstract

Individuals affected by inherited neuromuscular diseases often present with a specific pattern of muscle weakness, which can guide clinicians in genetic investigations and variant interpretation. Nonetheless, more than 50% of cases do not receive a genetic diagnosis. Oculopharyngodistal myopathy (OPDM) is an inherited myopathy manifesting with a particular combination of ptosis, dysphagia and distal weakness. Pathologically it is characterised by rimmed vacuoles and intranuclear inclusions on muscle biopsy. In recent years GCC • CCG repeat expansion in four different genes have been identified in individuals affected by OPDM in Asian populations. None of these have been identified in affected individuals of non-Asian ancestry.

In this study we describe the identification of CCG expansions in *ABCD3* in affected individuals across eight unrelated OPDM families of European ancestry. In two large Australian OPDM families, using a combination of linkage studies, short-read WGS and targeted ONT sequencing, we identified CCG expansions in the 5’UTR of *ABCD3*. Independently, the *ABCD3* CCG expansion was identified through the 100,000 Genomics England Genome Project in three individuals from two unrelated UK families diagnosed with OPDM. Targeted ONT sequencing confirmed the presence of mono-allelic CCG repeat expansions ranging from 118 to 694 repeats in all tested cases (*n*=19). The expansions were on average 1.9 times longer in affected females than affected males, and children of affected males were ∼2.3 times more likely to have the disease than those of affected females, suggesting inheritance of an expanded allele from an affected mother may have reduced penetrance. *ABCD3* transcripts appeared upregulated in skeletal muscle and cells derived from affected OPDM individuals, suggesting a potential role of over-expression of CCG repeat containing *ABCD3* transcript in progressive skeletal muscle degeneration. The study provides further evidence of the role of non-coding repeat expansions in unsolved neuromuscular diseases and strengthens the association between the GCC • CCG repeat motif and a specific pattern of muscle weakness with prominent cranial involvement across different populations.

## INTRODUCTION

Oculopharyngodistal myopathy (OPDM) was first delineated and described in four families presenting with autosomal dominant disease by Satoyoshi and Kinoshita in 1977.^1^ Individuals with OPDM typically present with adult-onset progressive ptosis, external ophthalmoplegia and weakness of the facial, masseter and pharyngeal muscles, resulting in dysphagia and weakness of the distal limb muscles. Muscle biopsies show chronic myopathic changes, including the presence of rimmed vacuoles and myeloid bodies within the myoplasm and intranuclear filamentous inclusions.^2^ These intranuclear inclusions are also evident in skin biopsies.^3^

It would be more than 40 years before the genetic aetiology of OPDM started to be untangled. To date, heterozygous GCC • CCG repeat expansions in the 5’UTR of four genes (*LRP12*,^4^ *GIPC1*,^5^ *NOTCH2NLC* ^6,7^ and *RILPL1*) have been associated with OPDM in Asian populations, either isolated or as part of a more complex neurological condition.^8,9^ However, as yet no OPDM case of European origin has been found to carry CGG repeat expansions at any of the previously identified OPDM loci.

This study sought to identify the genetic cause of the OPDM disease in multiple families of European ancestry. Herein we describe and characterise eight unrelated OPDM families, encompassing 35 affected individuals, and identify CCG expansions in the 5’UTR of *ABCD3* as the cause of cranial and distal limb weakness in all families. Additionally, we identified an increased expression of the CGG expansion containing *ABCD3* transcript, as a possible disease mechanism underlying muscle degeneration.

## METHODS

### Cohort

Individuals with OPDM and genetically unconfirmed myopathy were recruited from participating centres in Australia, the United Kingdom (UK), France and through a network of collaborators worldwide (OPDM study group). Available clinical and pathological data were reviewed.

### Genetic studies

Genetic testing performed in this cohort includes short read (sr) whole exome and whole genome sequencing (WES/WGS), linkage analysis and haplotype analysis. For repeat expansion identification, all samples were profiled with EHdn v0.9.0 to obtain a count of anchored in-repeat reads. RP-PCR was performed to study segregation of the identified CCG expansion in families and to screen additional cases who did not undergo srWES or srWGS. Targeted ONT sequencing and optical genome mapping were performed to confirm the presence and estimate the size of the CCG repeat expansion in all affected individuals with sufficient DNA or in fresh blood, when available, respectively.

### RNA and protein studies

*ABCD3* transcript expression was quantified by RNA-seq performed on RNA extracted from skeletal muscle and RNA *in situ* hybridisation on human derived skin fibroblasts, from individuals and controls.

The presence of intranuclear inclusions was tested by performing p62 staining on skin biopsies and fibroblasts, from affected individuals and controls.

Detailed methods are provided in the supplementary material.

## RESULTS

We report 35 OPDM individuals from eight unrelated families from Australia, the UK and France presenting with OPDM, including six with clear autosomal dominant inheritance and present detailed clinical information for 24 affected individuals (**Table 1**).

*Identification of CGG repeat expansion in the ABCD3 5’UTR in affected individuals with OPDM* Combined linkage performed for two of the families, AUS1 (n=5 affected individuals) and AUS2 (n=5 affected individuals), produced a maximum combined multipoint logarithm of odds (LOD) score of 2.98 for SNPs between and including rs12142220 to rs490680. This comprised a ∼24 Mb linkage region spanning chr1: 84,662,217 – 108,724,518, hg19). The LOD score of 2.98 was achieved at rs1801265. Analysis of srWGS from AUS1-V:3 using EHdn identified a CCG repeat expansion within the untranslated first exon of *ABCD3* and called this expansion as 72 repeats. Visual inspection of the BAM file in the Integrated Genome Viewer (IGV) revealed that the 5’UTR of *ABCD3* was spanned by >15 reads which confirmed the presence of a CCG repeat expansion (**Figure 1A**).

**Figure 1:**
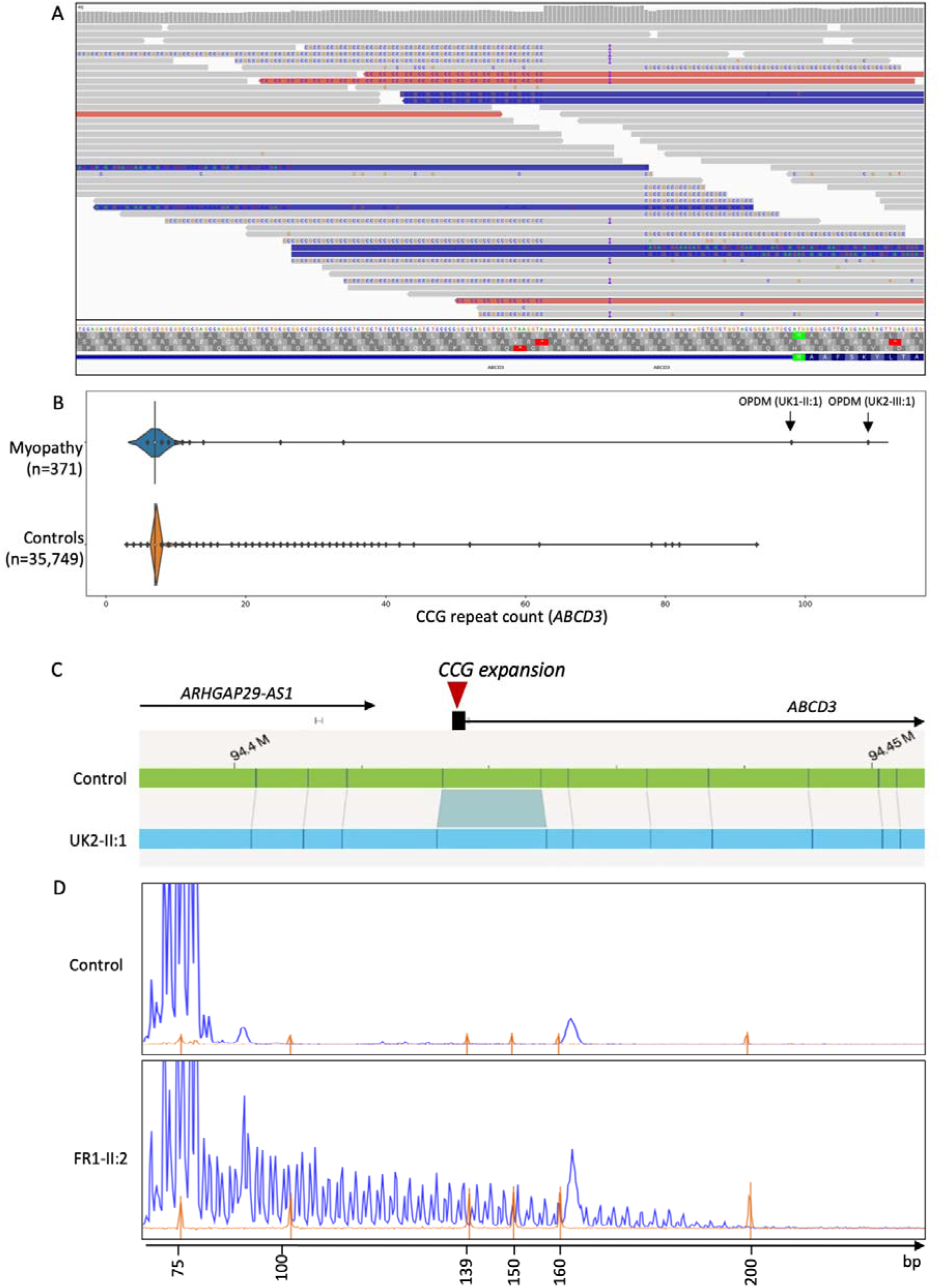
Identification of an expanded CCG repeat within the 5’UTR of ABCD3 in OPDM. (**A**) srWGS data (AUS1-V:3) in IGV showing ∼50% of reads mapped to the *ABCD3* promoter contain expanded repetitive sequences. (**B**) Identification of *ABCD3* CCG expansion causing OPDM in srWGS through Genomics England 100,000 Genome Project. (**C**) Outputs from the Bionano Access® Software showing the presence of an expansion within the 5’UTR of *ABCD3* in an OPDM individual relative to a healthy control. (**D**) Electropherogram of repeat-primed PCR. The upper panel showed no repeat expansion in unaffected individual while the lower panel shows a saw-tooth tail pattern of the repeat expansion in an affected OPDM individual (FR1-II:1).

Notably, the *ABCD3* repeat locus was independently identified through an unbiased analysis of the Genomics England 100,000 Genome Project – Rare Disease Cohort, thus strengthening the validity of the finding. Based on the presence of a shared pathogenic expanded CGG • CCG repeat motif in different genes underlying OPDM in cases of East Asian ancestry, we speculated that CGG • CCG expansion in additional genes may cause OPDM in cases of European origin. Therefore, we leveraged srWGS data from 378 cases with myopathy enrolled in the 100,000 Genome Project. We specifically looked for loci containing CGG • CCG expansions, which were enriched in the myopathy group compared to controls. The locus with the highest z-score identified by EHdn (outlier method) was a CCG repeat located in the 5’UTR of *ABCD3*. We next performed a more accurate profiling of the repeat locus using ExpansionHunter v3.2.2 which confirmed the presence of a large monoallelic CCG expansion of *ABCD3* in two cases diagnosed with OPDM (estimates of 109 and 98 repeats) (**Figure 1B**).

The CCG expansion in *ABCD3* was confirmed by RP-PCR and segregated with the disease in the family UK2, being present in the affected sister UK2-III:1 but absent in their unaffected mother. Unfortunately, DNAs from additional affected family members (UK2-II:1 and UK2-I:2) and the obligate carrier UK2-II:2 were unavailable for testing, as these individuals were deceased.

We next performed RP-PCR to screen the CCG expansion in *ABCD3* in a cohort of 68 cases diagnosed with cranial or distal limb myopathy or full OPDM. We identified three additional probands of French nationality from unrelated families diagnosed with OPDM (FR1-II:1, FR2-II:2, FR3-II:1) carrying the CCG expansion in *ABCD3* (**Figure 1D**). In UK1-II:1, UK2-III:1, UK2-III:2 and FR1-II:1 fresh blood was available for extraction of ultralong fragments. Therefore, these samples were subjected to Bionano optical genome mapping, confirming the presence of the *ABCD3* repeat expansion (**Figure 1C**).

The existence of two obligate carriers (AUS2-III:4 and UK2-II:2), who both lived until their old age and did not develop the disease, suggests that the repeat may show reduced penetrance in rare cases. Unfortunately, DNA was not available for assessing the repeat size.

*ABCD3* repeat expansions were absent from optical genome mapping control datasets comprising 974 alleles and are very rare in large srWGS control datasets. There were no large expansions over

50 repeats, corresponding to the average read length of 150bp, in 32,884 alleles present on GnomAD. Only one expanded allele out of 6,538 (0.015%) was identified in the Broad Centre for Mendelian Genomics and Rare Genomes Project cohorts and ten expanded alleles out of 71,498 alleles (0.013%) were identified in the 100,000 Genomes Project. In all control individuals the estimated repeat size was lower compared to OPDM cases. Further details of repeat size distribution in control datasets are available in **Supplementary results and Supplementary Figure S1**.

### Targeted OPDM ONT sequencing accurately sizes ABCD3 repeat expansions and identifies that the locus remains unmethylated in most OPDM individuals

Targeted long-read sequencing (LRS) of the *ABCD3* 5’UTR was performed on DNA from ten affected and two unaffected individuals from families AUS1-3 and nine affected individuals from families UK1, UK2, FR1, FR2 and FR3. LRS identified pure CCG repeat expansions on a single allele in all affected individuals (*n*=19) ranging from 118 to 694 repeats (average 283 repeats, SEM: 39.8). In all affected individuals the second (short) allele contained seven CCG repeats, the unaffected relatives harboured two alleles containing seven CCG repeats (**Figure 2A**). The expanded alleles remained unmethylated in 17/19 OPDM individuals, while in two affected females harbouring large expansions of the expanded allele was hypermethylated (**Supplementary Figure S2**). Anecdotally, FR3-I:2 had the largest expansion of 624 repeats, but age of disease onset was relatively late, in her fifth decade of life, suggesting that the hypermethylation of very large repeat could mitigate the phenotype or reduce its penetrance.

**Figure 2:**
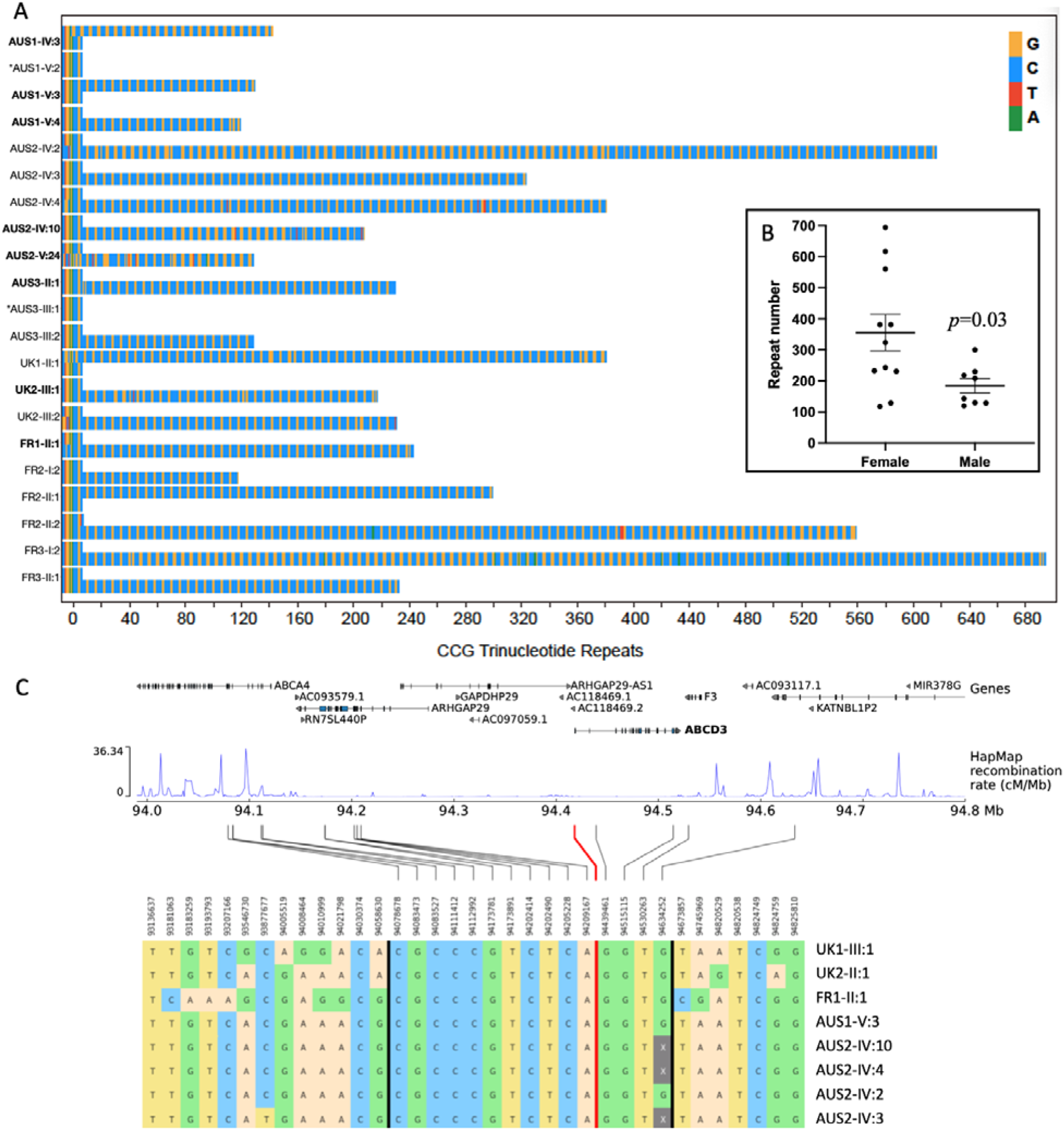
Targeted long-read sequencing and analysis of the ABCD3 repeat expansion. (**A**) Long-read genotyping of the *ABCD3* 5’UTR STR expansion showed heterozygous pure CCG expansions in all affected individuals (*n*=19). The wild-type alleles in affected individuals and two unaffected relatives contained 7 x CCG repeats. Unaffected individuals are denoted by an *. Individual IDs for affected males are bolded. (**B**) Repeat expansions are larger and more variable in size in affected females compared to affected males. (**C**) The shared haplotype region of 560 kb, identified by 15 SNPs between the vertical black bars. The red line indicates the position of the *ABCD3* repeat expansion. Haplotypes were obtained by phasing genotype data (WGS and WES) with SHAPEITv4. Since the rightmost SNP in the shared haplotype is only available for WGS samples, the four WES samples have been greyed out.

No repeat expansions at the previously reported loci (*LRP12*, *GIPC1*, *NOTCH2NLC, RILPL1*) were identified in any affected individuals, nor were any other alternate candidate repeat expansions identified by EHdn that were shared by the majority of the affected individuals.

### Effect of sex on repeat size and transmission

There was a negative correlation between repeat expansion size and age-of-onset in affected males (*y*=3.029×+272.8, *n*=6, *p*=0.0063) with larger expansions associated with earlier onset of disease. The repeat expansion size in affected males was 185±23 repeats (mean±SEM, *n*=8) with a range of 180 repeats; in affected females, the repeats were ∼1.9 times longer (356±59 repeats, *n*=11, *p*=0.0295) and more variable in length (range 576 repeats) **Figure 2B**, suggesting the expansion may be more unstable in females. The onset of the disease in affected females was typically ∼5 years earlier than in affected males (24±2.9 vs 29.8±4.9) without a significant correlation with repeat expansion size (**Supplementary Figure S3)**.

Additionally, maternal inheritance of the repeat appears to be associated with reduced penetrance in the offspring. Nine affected/obligate carrier males had 36 children, of which 19 were also affected (penetrance 52.8%), while 11 affected females had 31 children, but only seven were affected (penetrance 22.6%) (Chi square 6.4, df =1, *p*=.011). Also, the two obligate carriers AUS2-III:4 and UK2-II:2 appeared to have inherited the expanded allele from an affected mother.

### All individuals carrying the ABCD3 CCG expansion share an ancestral haplotype

Subsequently, we looked at the inferred haplotypes associated with *ABCD3* CCG expansions. We identified a region of 450 kb which was shared among all *ABCD3* CCG OPDM samples (14 SNPs between chr1:94078678-94530263), encompassing the entire *ABCD3* gene (**Figure 2C**). An additional SNP at position chr1:94634252, available only for samples with srWGS data, extended the shared region by an additional 110 kb downstream of *ABCD3* (total 560 kb). The shared haplotype lies primarily within a low-recombination region (HapMap data) and has an allele frequency of 0.2% in the 100,000 Genome Project – Rare Disease Cohort (myopathy cases excluded, AN=27,684).

### Individuals carrying the CCG ABCD3 repeat expansion present with the hallmark features of OPDM

Comprehensive clinical data from 24 individuals carrying CCG *ABCD3* expansions were available for review (**Table 1**). The average age of onset was 26.7 years (range: 10-50 years). There were 11 affected males (average age at onset 29.8 years) and 13 affected females (age of onset 24.0 years). In almost all cases the presenting symptom was ptosis (22/24, 92%); one presented with dysphagia and one with weakness of the oropharyngeal muscles. On examination, ptosis was confirmed in all affected individuals, while other common features included ophthalmoparesis (14/20, 70%), facial weakness (16/21, 77%), dysphagia (16/21, 77%), dysarthria (13/20, 65%) and distal lower limb weakness (17/23, 74%). Nerve conduction studies and electromyography were in keeping with a myopathic process. CK was normal to mildly elevated (278±79 IU, range 142-377).

Skeletal muscle biopsies from nine affected individuals were available for review. Light microscopy revealed the hallmark feature of rimmed vacuoles and other features such as numerous internal nuclei, myofibre splitting, increased fibrous tissue, and internalised capillaries. Rimmed inclusions stained positive for AMP deaminase (**Figure 3**). Autophagic vacuoles and myeloid bodies were evident on electron microscopy. Rare intranuclear p62-positive inclusion were identified only in one out of five cases. Muscle biopsies from two cases were available for re-imaging with electron microscopy; careful examination of these biopsies did not identify any intranuclear inclusions (data not shown). It is noted that intra-myonuclear inclusions are only present in ∼1% of nuclei in other genetic forms of OPDM.^10^

**Figure 3:**
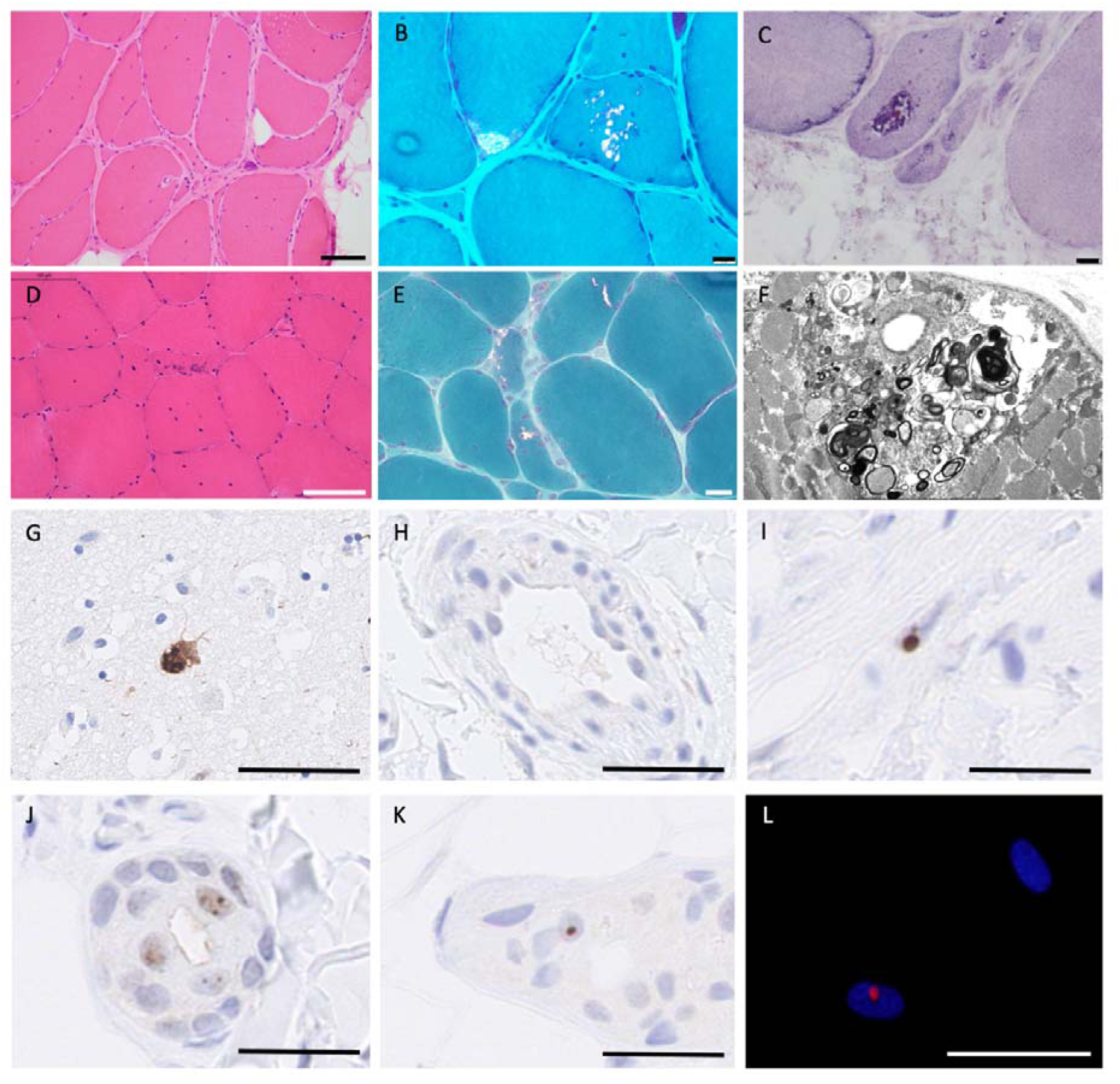
Muscle pathology in ABCD3-related OPDM. Haematoxylin and eosin staining (**A, D**) and modified Gomori trichrome staining (**B, E**) show rimmed vacuoles, internal nuclei, variation in myofibre size and myofibre splitting. Increased fibrotic tissue is also evident. Inclusions also stain with AMP deaminase (**C**). Subsarcolemmal myeloid bodies and autophagic vacuoles are evident on electron microscopy (**F**). (**A-C, F**) AUS2-IV:2, (**D**) AUS3-II:1 and (**E**) AUS1-III:3. p62 staining (**G-L**) showed intranuclear aggregates in fibroblasts (**I**), and exocrine glands (**J, K**) from OPDM individuals UK1-II:1 and UK2-III:2 but not in control skin (**H**). FFPE slide from post-mortem brain from an individual with dementia showing a strong p62 neuronal intranuclear signal used as positive control for the staining (**G**). (**M**) A p62-positive (red) intra-nuclear inclusion in cultured primary skin fibroblast from an OPDM individual (AUS1-IV:3). Scale bars: 20 µm (B, C, E), 50 µm (A, H, I, J, K, L, M), 100 µm (A, D, G).

As intranuclear inclusions can often be observed in the skin of individuals carrying CCG repeat expansions, including neuronal intranuclear inclusion disease, we performed immunohistochemistry in two individuals with OPDM and carrying *ABCD3* expansions. Intranuclear p62-positive inclusions were identified in nuclei of exocrine glands, keratinocytes and fibroblasts in individuals’ biopsies (**Figure 3H-K**) but were absent or less prominent in controls. Staining of primary fibroblasts generated from skin biopsy of AUS3-IV:3 for p62 found ultra-rare intranuclear p62-positive inclusions (<0.1%, **Figure 3L**). Overall, p62-positive nuclei were rare (<1%) in the skin.

### Investigations of disease mechanisms

RNA sequencing and analysis from affected muscle tissue, based on normalised gene counts calculated by OUTRIDER, found that *ABCD3* expression in the three OPDM individuals was higher compared to cases affected by other neuromuscular diseases and healthy controls (**Figure 4A**). Skeletal muscle from AUS3-II:1 had the highest expression (2,030 [normalised gene count]), followed by AUS2-IV:4 (1,582), and AUS2-IV:2 (1,404). *ABCD3* was detected as an over-expression outlier in AUS3-II:1 (adjusted *p-*value = 1.8 x 10^-3^ log2-fold change = 0.88), but not in AUS2-IV:4, or AUS2-IV:2.

**Figure 4:**
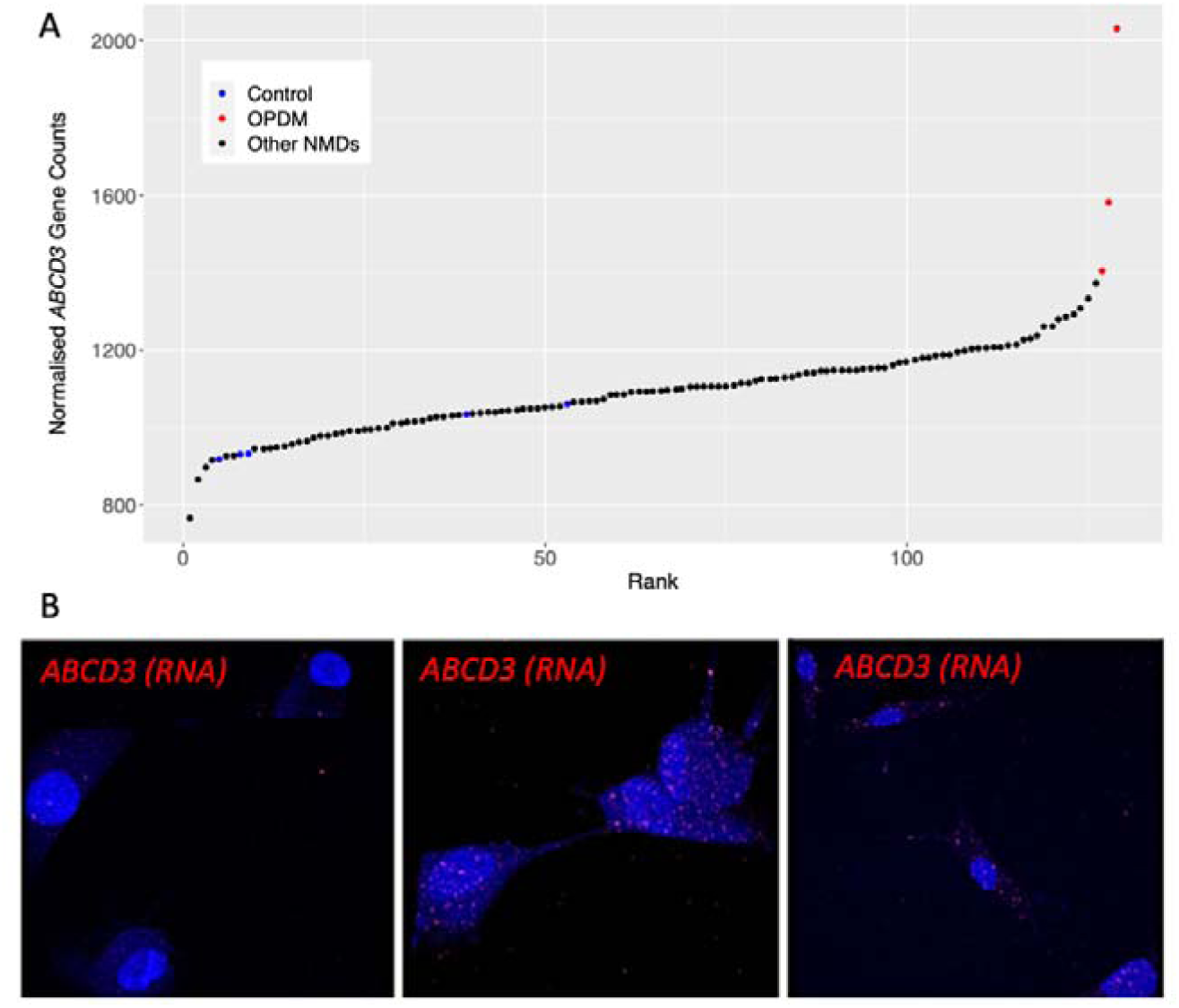
ABCD3 is over-expressed in OPDM skeletal muscle and forms nuclear and cytoplasmic foci in OPDM-derived fibroblasts. Analysis of skeletal muscle RNA-seq showed that *ABCD3* appears to be over-expressed in OPDM skeletal muscle compared to healthy controls (**A**). The normalised counts were obtained from the OUTRIDER results table. Detection of *ABCD3* transcript in fibroblasts by HCR-RNA FISH. In the OPDM line, the intensity of signal (in red) is increased and tends to aggregate in nuclear and cytoplasmic foci (**B**).

A key pathological hallmark of repeat expansion disorders is the presence of repeat-containing RNA foci. Therefore, we next performed HCR™ RNA-FISH for the *ABCD3* sense transcript on skin fibroblasts from one individual (UK2-III:1) and three age-matched controls. Compared to controls, we identified increased cytoplasmic and intranuclear signal of *ABCD3* transcript in individual-derived fibroblasts (**Figure 4B**). This is congruent with increased *ABCD3* transcript expression detected in skeletal muscle RNA-seq. In some fibroblasts, the *ABCD3* signal appeared more clustered to form foci in nuclei and cytoplasm. *ABCD3-*positive foci were identified in the skin biopsy of one OPDM individual but were exceedingly rare (one out of >100 nuclei, **Supplementary Figure S4**). No foci were identified in the skeletal muscle biopsy.

## DISCUSSION

In this study we describe the identification of CCG expansion in 5’UTR of *ABCD3* causing OPDM in Europeans. A combination of linkage analysis in two large Australian OPDM families (of British origin) and STR analysis of srWGS using EHdn identified the *ABCD3* repeat expansion as a candidate for disease in these families. Independently, analysis using EHdn on the Genomics England data identified two unrelated individuals within a neuromuscular disease cohort with a clinical diagnosis of OPDM with the same *ABCD3* expansion as outliers. Screening by RP-PCR of additional probands of European ancestry with cranial and distal limb weakness identified four additional unrelated OPDM families (three French and one Australian) with expansions in *ABCD3*.

European individuals with CCG *ABCD3*-related OPDM showed similar features to previously reported East Asian individuals with OPDM.^9,11^ Onset of the disease was usually in the third decade of life, although the range was broader from the 1^st^ to the 6^th^ decade, and ptosis was usually the presenting symptom. Indeed, upon examination, all individuals had ptosis, while ophthalmoparesis, facial weakness, pharyngeal weakness and distal weakness were present in ∼70%. In two families there are two male carriers who lived until old age who did not manifest the disease; unfortunately, we have not been able to size the expanded allele in these individuals. Muscle biopsy often showed mild myopathic changes with rimmed vacuoles, while intranuclear inclusions were exceedingly rare. This aligns with previous reports in East Asian OPDM individuals where intranuclear inclusions were identified in less than 1% of the nuclei in affected muscle tissue.^10^

Interestingly, all affected European individuals shared a 560 kb ancestral haplotype encompassing *ABCD3,* which has an allele frequency of 0.2% in the Genomics England 100,000 Genomes Project. There are rare occurrences *ABCD3* repeat expansions (>50 repeats, *n*=11 alleles) being detected in srWGS data from individuals without OPDM in both the Genomics England and gnomAD datasets (<∼0.01%). In two such individuals within Genomics England, for whom follow-up was possible, targeted ONT sequencing found the repeat expansions to be 88 and 120 repeats, below the pathogenic range observed in this OPDM cohort. Of note, Ishiura *et al.* also found CGG repeat expansions in *LRP12* in a small number of control subjects (0.2%). Also, hexanucleotide repeat expansions in *C9orf72* that cause ALS are seen in 0.15% of UK controls^12^ and 0.4% of Finnish controls.^13^ Thus, rare occurrences of repeat expansions in healthy individuals are a recurring finding in noncoding repeat expansion diseases, including other genetic forms of OPDM, and suggest the possibility of additional factors, including so far unknown genetic modifiers, contributing to disease penetrance and expressivity.

Our study revealed that *ABCD3* expansion accounted for a relevant proportion of OPDM cases in the Caucasian population, with 15 cases (62%) exhibiting a complete OPDM phenotype. This indicates that *ABCD3* CCG expansions should be considered when investigating individuals with isolated weakness of ocular, facial bulbar or distal. Notably, over 70 OPDM or OPDM-like European probands tested in this study remain genetically unexplained, indicating the presence of further genetic heterogeneity underlying this condition, even within the European population. While *GIPC1*, *RILPL1*, *NOTCH2NLC* and *LRP12* accounted for ∼75% of Asian individuals with OPDM,^12^ we did not identify any individuals with expansions in these four genes within our European OPDM cohort.

Targeted long-read sequencing of 19 *ABCD3* OPDM individuals from eight families found that the pathogenic range for *ABCD3* repeats is 118-694, with repeats being ∼1.9 times larger in affected females than males. Although there is no definite explanation for this observation it is possible that the CCG *ABCD3* repeat is prone to further expansion in female embryos. Of note, this phenomenon has been reported in Huntington disease^14,15^ and it is hypothesised that X– or Y-chromosome encoded factors involved in DNA replication or repair may influence the repeat stability during the early stages of embryo development.^15^

Despite the larger repeat size in females *ABCD3* OPDM individuals were much more likely to inherit the disease from an affected father than an affected mother. We speculate that large maternally inherited *ABCD3* expansions may be prone to further expansion resulting in hypermethylation and silencing of the expanded allele. However, we cannot rule out that very large expansions (>700 repeats) would be deleterious to oocytes or embryonically lethal. Zeng *et al.* reported that contractions of ultralong *RILPL1* repeats were more common in male-to-offspring transmission than female-to-offspring transmission.^9^ This parent-of-origin effect, which is common to several repeat expansion diseases, should be taken into account when counselling affected individuals and their families, as male individuals carrying CCG *ABCD3* repeat expansion are at higher risk of having affected children.

The identification of GCC • CGG repeats as the shared genetic cause of OPDM in functionally different genes suggests a shared pathogenic mechanism underlying myo– and neurodegeneration, at least partly independent of the repeat-containing genes.^8,16^ Interestingly, there appears to be a relatively confined pathogenic range associated with GCC • CCG expansions, which is different from most other repeat expansion diseases. Indeed, both small and very large GCC • CCG expansions seem to be tolerated, while disease-causing expansions occur within a specific window.

Although the mechanism underlying muscle degeneration in *ABCD3* repeat expansion remains unknown, RNA-sequencing found that *ABCD3* is over-expressed in skeletal muscle from *ABCD3* OPDM individuals. HCR RNA-FISH for *ABCD3* sense transcripts also confirmed an increased transcript abundance in patients’ fibroblasts compared to controls, with clusters of signal in some nuclei suggestive of RNA foci. These foci are thought to represent repeat-containing RNAs and specific RNA-associated binding proteins, which could be detrimental to myofibre survival, a mechanism that is well documented in myotonic dystrophy^17^ and suggested in another OPDM subtype.^8,9^ Potential RNA toxicity mechanisms have also been proposed in *RILPL1*-related OPDM.^9^ Alternatively, or concurrently, repeat peptides which are synthesised through repeat associated non-ATG dependent (RAN) translation of CGG repeats, as shown in *NOTCH2NLC*-related neuronal intranuclear inclusion disease, could contribute to the progressive muscle degeneration observed in *ABCD3*-mediated disease. Using a recently developed *in-silico* tool^18^ we predicted that the most likely amino-acid stretches produced from RAN translation of the *ABCD3* repeat expansion are alanine (sense) and alanine or glycine (anti-sense) (**Supplementary Figure S4**). Experimental determination of the contribution of RAN-mediated translation to the pathobiology of *ABCD3* repeated OPDM is beyond the scope of this initial study.

In summary, we describe here the identification of CCG repeats in the 5’UTR of *ABCD3* as a cause of OPDM in families of European ancestry and show that all affected individuals share a common ancestral haplotype. This study improves our understanding of the molecular aetiology of OPDM and suggests that CCG and CGG expansions in 5’UTRs of other genes may explain additional cases of OPDM.

## Data Availability

All data produced in the present study are available upon reasonable request to the authors

## ACKNOWLEDGEMENTS

We thank Profs Ichizo Nishino, Carolin Sewry, and Werner Stenzel for reading and review of the muscle biopsies and electron microscopy.

This project was supported by an NHMRC Ideas Grant (APP2002640) to G.R, N.G.L, M.R.D., P.J.Lamont and M.C-S and Medical Research Council (MR/T001712/1), Fondazione Cariplo (grant n. 2019-1836), the Inherited Neuropathy Consortium, Fondazione Regionale per la Ricerca Biomedica (Regione Lombardia, project ID 1751723) and Italian Ministry of Health (Ricerca Corrente 2021-2022) to A.C. C.K.S. and L.D. are supported by an Australian Government Research Training Program (RTP) Scholarship, I.W.D. is supported by an MRFF Investigator Grant (MRF1173594) and G.R. is supported by an NHMRC EL2 Investigator Grant (APP2007769). M.B. is supported by an NHMRC L1 Investigator Grant (APP1195236), This work was also supported by the Australian State of Victoria’s Government’s Operational Infrastructure Support Program, and the NHMRC Independent Research Institute Infrastructure Support Scheme (IRIISS). This work was supported by resources provided by the Pawsey Supercomputing Research Centre with funding from the Australian Government and the Government of Western Australia. Library preparation and RNA-sequencing was conducted in the Genomics WA Laboratory in Perth, Australia. This facility is supported by BioPlatforms Australia, State Government Western Australia, Australian Cancer Research Foundation, Cancer Research Trust, Harry Perkins Institute of Medical Research, Telethon Kids Institute and the University of Western Australia. We gratefully acknowledge the Australian Cancer Research Foundation and the Centre for Advanced Cancer Genomics for making available Illumina Sequencers for the use of Genomics WA.

## DECLARATIONS

H.G. has previously received travel and accommodation expenses from ONT to speak at conferences. H.G. and I.W.D. have paid consultant roles with Sequin PTY LTD. MCF has paid consultancy roles with Fenix Innovations, the Victorian Government and PTC. The other authors have no conflicts of interest to declare.

## SUPPLEMENTARY METHODS

### Patients

Patients diagnosed clinically with OPDM and genetically unconfirmed myopathy were recruited from participating centres in Australia, the United Kingdom (UK), France and through a network of collaborators worldwide (OPDM study group). The following information was collected for all patients using a standardised template: demographics, family history, current age, age at disease onset, first symptom/s, presence of ptosis, ophthalmoparesis, facial weakness, dysphagia, dysarthria, distribution of limb weakness, respiratory involvement, age at death. When available, EMG were reviewed.

Muscle biopsies were performed on nine individuals and routine histopathology conducted as part of the clinical work-up. Investigations included H&E, Gomori trichrome and NADH staining. P62 staining was also available in X cases. Ultrastructural studies were performed following standard methods.^1^ A small fragment of the muscle was fixed in 2.5% w/v glutaraldehyde solution, postfixed in 1% w/v osmium tetroxide, and embedded in epoxy resin. Semithin sections were stained with 1% toluidine blue. Ultrathin sections were mounted on copper grids and examined with a Zeiss Libra 120 transmission electron microscope.

This study was approved by the Human Research Ethics Committee of the University of Western Australia (RA/4/20/1008), the Human Research Ethics Committee of the Royal Children’s Hospital (HREC 28097) and Northeast-Newcastle & North Tyneside 1 Research Ethics Committee (22/NE/0080). All individuals provided informed consent and we obtained consent from the affected individuals or their relatives (in the case of deceased individuals) to share clinical images.

### Linkage

Linkage was performed using MERLIN^2^ and whole exome sequencing data (WES) on five affected individuals from AUS1 (III:3, IV:3, IV:4, V:3, V:4) and four affected individuals from AUS2 (IV:2, IV:3, IV:4, IV:10).

### Short read (sr)-WES and srWGS and analysis in Australian families

Ampliseq srWES was performed on DNA from affected individuals in the Australian families (AUS1, AUS2) as described previously.^3^

PCR free srWGS (2 x 150 bp reads, >30× coverage) on the proband in Family AUS1 (V:3) was performed at The Center of Mendelian Genomics, The Broad Institute. ExpansionHunter denovo (EHdn)^4^ was used to identify repeat expansions within the linkage region.

### srWGS analysis in the Genomics England 100,000 Genome Project

The 100,000 Genomes Project, run by Genomics England, was established to sequence whole genomes of patients of the National Health Service (NHS) of the UK, affected by rare diseases and cancer.^5^ We analysed a cohort of 371 myopathy cases and 35,749 non-neurological controls enrolled in the 100,000 Genome Project – Rare Disease Cohort. For all individuals srWGS was available (paired-ended, 150bp, average read depth > 30×). All samples were profiled with EHdn v0.9.0 to obtain a count of anchored in-repeat reads, then jointly analysed using the “outlier” method provided in the EHdn package. The method computes a z-score for each sample, based on the original distribution of read counts. The score is to be interpreted as distance to the median, relative to the distribution width.

### ABCD3 haplotype analysis

We used SHAPEITv4 with default parameters to phase a 3Mb region (chr1:92500000-95500000 Hg38) encompassing *ABCD3*. To maximise available haplotype information, the entire Rare Diseases cohort of Genomics England (78,195 samples from patients affected by rare diseases, including the OPDM cases UK1-II:1 and UK2-III:2) was jointly phased. Input data format was an aggregate VCF file with a total of 599,962 variants. External OPDM cases (AUS1-V:3, AUS2-IV:2, AUS2-IV:3, AUS2-IV:4, AUS2-IV:10, FR1-II:1) were subsequently phased using the previously phased data as reference.

### Repeat primed PCR (RP-PCR)

RP-PCR was performed to study segregation of CCG expansion in families UK1 and UK2 and to screen *ABCD3* CCG expansion in a cohort of 68 cases diagnosed with either OPDM (n=28), chronic progressive external ophthalmoplegia (n=25) either isolated or as part of more widespread myopathic involvement, or distal myopathy (n=15). The following primers were used: *ABCD3* Fw: FAM-GCTCTCCTCCCAGTCTCCCC; anchor: CAGGAAACAGCTATGACC; *ABCD3* Rv: CAGGAAACAGCTATGACCCGGCGGCGGCGG. The PCR mix contained 0.5 U Takara LaTaq (Takara, Shiga, Japan), 1X GC Buffer, 400 μM each dNTP mixture, 0.4 μM primer *ABCD3*-Fw, anchor and 0.04 μM *ABCD3*-Rv, <1 μg DNA, and ddH2O for a final volume of 50 μL. We used the following thermal conditions: pre-denaturation (95 °C for 5 minutes), followed by 50 cycles of 95 °C for 30 seconds, 62 °C for 1 minute, 72 °C for 2 minutes and final extension (72 °C for 5 minutes). The ramp rate to 95 °C and 72 °C was set to 2.3 °C.s^−1^ and that to 62 °C was set to 1.5 °C.s^−1^. Fragment analysis was conducted on an ABI-3730XL Sequencer, and data were visualised using Geneious Prime (Biomatters Ltd).

### Analysis of the ABCD3 locus in srWGS from population datasets

To examine the population distribution of STR alleles at the *ABCD3* locus, we ran ExpansionHunter on 16,442 srWGS samples from gnomAD by specifying the locus region as chr1:94418421-94418442 (GRCh38, 0-based coordinates) and repeat unit as GCC. We also used ExpansionHunter to genotype 3,270 srWGS rare disease samples from the Rare Genomes Project (RGP) and the Center for Mendelian Genomics (CMG). These included 1,608 cases from a wide range of disease and phenotype categories and 1,662 unaffected family members.

Finally, we profiled the *ABCD3* CCG expansion locus with EH in the entire 100,000 Genome Project cohort of 35,749 srWGS from non-neurological controls from the UK. We analysed its size in 724 controls, and 250 alleles from internal non-OPDM samples that underwent optical genome mapping at UCL Institute of Neurology (see methods below).

### Programmable targeted nanopore sequencing and analysis

High molecular weight (HMW) DNA samples were transferred to the Garvan Institute’s Sequencing Platform for long-read sequencing analysis on Oxford Nanopore Technologies (ONT) instruments. Prior to ONT library preparations, the DNA was sheared to ∼20kb fragment size using Covaris G-tubes and visualised, post-shearing, on an Agilent TapeStation. Sequencing libraries were prepared from ∼3-5ug of HMW DNA, using native library prep kit SQK-LSK110, according to the manufacturer’s instructions. Each library was loaded onto a FLO-MIN106D (R9.4.1) flow cell and run on an ONT MinION device with live target selection/rejection executed by the *ReadFish* software package.^6^ Detailed descriptions of software and hardware configurations used for *ReadFish* experiments are provided in a recent publication that demonstrates the suitability of this approach for profiling tandem repeats.^7^ Samples were run for a maximum duration of 72 hours, with nuclease flushes and library reloading performed at approximately 24– and 48-hour timepoints for targeted sequencing runs, to maximise sequencing yield.

Raw ONT sequencing data was converted to BLOW5 format^8^ using *slow5tools* (v0.3.0)^9^ then base-called using *Guppy* (v6). Resulting FASTQ files were aligned to the *hg38* reference genome using *minimap2* (v2.14-r883).^10^ The short-tandem repeat site within *ABCD3* was genotyped using a process validated in our recent manuscript.^7^ This method involves local haplotype-aware assembly of ONT reads spanning a given STR site and annotation of STR size, motif and other summary statistics using Tandem Repeats Finder^11^ followed by manual inspection and motif counting. DNA methylation profiling was performed with F5C (v1.1).^12^

Targeted ONT sequencing of the four previously described OPDM loci was also performed on affected individuals from each family.

### Bionano optical genomic mapping

Patients UK1-II:1, UK2-III:1, UK2-III:2, UK1-II:1 and FR1-II:1 for whom whole blood was available were subjected to Bionano optical mapping to gather additional information on the precise size of the expanded repeat. HMW genomic DNA was isolated using kits provided by Bionano Genomics, as described in Bionano Prep SP Frozen Human Blood DNA Isolation Protocol v2. Homogeneous HMW DNA was labelled using Bionano Prep Direct Label and Stain (DLS) Protocol with kit provided, and the homogeneous labelled DNA was loaded onto a Saphyr chip. Optical mapping was performed at theoretical coverage of 400×. Molecule files (.bnx) were then aligned to hg38 with Bionano Solve script “align_bnx_to_cmap.py” using standard parameters.

### Skeletal muscle RNA-seq

RNA was extracted from patient and control skeletal muscle biopsies (∼15-50 mg) using the RNeasy Fibrous Tissue Mini Kit (QIAGEN #74704). RNA purity was assessed by Nanodrop, followed by PCR amplification and 1% agarose gel electrophoresis to confirm the absence of DNA. Sample sequencing was performed at Genomics WA (Harry Perkins Institute of Medical Research, Perth, Australia). Stranded Poly A RNAseq libraries were prepared from extracted RNA using Agilent Sureselect XT library preparation kit. The protocol includes poly A enrichment followed by fragmentation, reverse transcription, ligation with adaptors and amplification for indexing. QC was performed using TapeStation 4200 and Qubit, and QC sequencing on the Illumina iSeq 100 flow cell. Paired-end sequencing (strand-specific reverse) was performed on the Illumina NovaSeq 6000 instrument using a 2 x 150 cycle configuration to a depth of 50 million read pairs per sample. Adaptor sequences were removed and demultiplexed FASTQ files were provided by Genomics WA for download and further analysis.

Processing of FASTQ files, including read quality control and alignment, was performed using the nf-core/rnaseq pipeline (https://nf-co.re/rnaseq/1.3), version 3.8.1. Raw reads were aligned to the GRCh38 human reference genome using STAR (version 2.7.10a). Detection of aberrant expression was achieved using DROP (version 1.3.3),^13^ as previously described.^14^ DROP leverages OUTRIDER,^15^ a method for detecting aberrant RNAseq read counts, which uses a denoising autoencoder to control for latent effects and returns multiple-testing correctedL*p*-values (FDR) for each gene and sample. The cohort consisted of 130 skeletal muscle RNA-seq datasets from rare neuromuscular disease patients, including three OPDM patients and five unaffected controls.

### Fibroblasts cultures

Human derived skin fibroblasts from OPDM patients (UK2-III:1 and AUS1-IV:3) and age-matched controls were cultured in Dulbecco’s Modified Eagle Media (DMEM) supplemented with 10% FBS at 37 °C in a 5% CO_2_ humidified incubator.

### Immunofluorescence and imaging

For detection of p62 aggregates in skin, formalin-fixed paraffin-embedded (FFPE) slides were stained with the mouse monoclonal antibody against p62 (1:100 dilution, 3/P62LCK Ligand, BD Transduction) and developed using the OptiView DAB IHC Detection Kit (Roche Diagnostics).

Primary fibroblast cultures from AUS1-IV:3 were grown in 24 well tissue culture plates, fixed and permeabilised with 2% paraformaldehyde and 1% saponin in phosphate buffered saline (PBS) for 15 minutes. Cells were then blocked in PBS containing 10% fetal calf serum, 5% goat serum and 1% bovine serum albumin (blocking buffer) for 60 minutes at room temperature. Cells were then incubated with a mouse monoclonal antibody against p62 (1:50 dilution, Abcam, ab56416) overnight at 4 °C. Cells were washed 3×5 minutes with PBS and then incubated with goat anti-mouse IgG2a AlexaFluor® 555 (1:500) for 60 minutes at room temperature in blocking buffer. Cells were washed in PBS and counterstained with Hoechst (Sigma, Australia). Imaging was performed on an inverted fluorescent microscope (model IX-71, Olympus) with a digital camera (model DP-74, Olympus).

### Hybridisation Chain Reaction (HCR) RNA Fluorescence in situ hybridisation (FISH)

We performed RNA *in situ* hybridisation for *ABCD3* transcript on human derived skin fibroblasts from one patient (UK2-III:1) and three age-matched controls. HCR™RNA-FISH significantly amplifies the signal of an individual molecule over traditional FISH and was shown to be more sensitive than FISH at detecting low abundant RNAs and endogenous G4C2 repeats in *C9orf72* ALS-FTD patient brains.^16^ HCR™RNA-FISH (Molecular Instruments) was performed on fibroblasts according to manufacturer’s protocol. Briefly, 4×10^5^ cells were seeded on coverslips and fixed in 4% formaldehyde for 10 minutes at room temperature. Cells were then washed twice in PBS and permeabilised in 70% cold ethanol overnight at –20 °C. After two washes in 2× saline sodium citrate (SSC), cells were pre-warmed in probe hybridisation buffer (Molecular Instruments) at 37 °C for 30 minutes and then incubated with 1.2 pmol of *ABCD3* probe set in hybridisation buffer at 37°C overnight. The following day cells were washed four times with probe wash buffer (Molecular Instruments) at 37 °C and twice with 5× SSC + 0.1% Tween-20 at room temperature (5× SSC-T). A pre-amplification step was performed with probe amplification buffer (Molecular Instruments) for 30 minutes at room temperature and cells were incubated with 18 pmol hairpins B1H1 and B1H2 at room temperature overnight. Cells were washed 5× SSC-T at room temperature, incubated with DAPI for 15 minutes and then mounted with Dako mounting medium.

Images were taken on a Zeiss LSM 710 confocal microscope at 40× magnification. The signal was normalised to control samples.

### Statistical analyses

Data on *ABCD*3 repeat length are presented as mean +/+ standard error of the mean. A two-tailed t-test was conducted to determine if there was a significant difference between *ABCD3* repeat expansion size in affected females compared to affected males, with a threshold for significance set at *p*<0.05. To compare the proportion of affected children born to affected females vs affected males a Chi square test was performed, with significance set at *p*<0.05.

## SUPPLEMENTARY RESULTS

### ABCD3 repeat expansions are very rare in srWGS and optical genome mapping control datasets

Analysis of 16,442 gnomAD samples using ExpansionHunter found that the repeat size distribution had a mean of 7.1 x CCG repeats and a median of 7 x CCG repeats. The median is also 7 x CCG for each gnomAD subpopulation (ASJ, AMI, AMR, MID, SAS, EAS, NFE, FIN, AFR). The most expanded genotype among the gnomAD samples was 7/44 (CI: 7-7/43-58). The REViewer read visualisation for this sample contained several low-quality read alignments but supported a genotype of at least 38 repeats in the long allele, implying that ExpansionHunter works reasonably well for expansions at this locus **(Figure S1)**.

Two samples in the CMG and RGP cohorts (n=3,270) had expansions longer than any of those found in gnomAD (ie. > 44xCCG). The first sample was an unaffected father of an affected daughter with holoprosencephaly and hearing loss. ExpansionHunter reported his genotype as 7/81 (CI: 7-7/64-105). The other sample is the proband from OPDM family AUS1-V:3 and had a genotype of 7/80 (CI: 7-7/63-106). We have not been able to ascertain any further clinical details for the gnomAD individual with at least 38 repeats or the unaffected father in the RGP cohort). Thus, we do not know if they have an OPDM phenotype.

Analysis of 35,749 WGS from non-neurological controls enrolled in the Genomics England 100,000 Genome Project showed a median size for *ABCD3* repeat of 7 repeats and mean 7.17. EH estimated ten out of 71,498 alleles (0.013%) to have expansions >50 CCG repeat, corresponding to the average read length of 150bp. In all, the estimated repeat number was lower than the two OPDM cases (estimated repeat sizes: 52, 52, 52, 52, 62, 78, 80, 81, 82 and 93 repeats; **Figure 1B**). Genomics England policy does not allow for contact and examination of reportedly unaffected individuals; therefore, we could not obtain more DNA for precise sizing of the repeat or access medical files for these 10 subjects. We performed a similar analysis in the complete cohort of 14,600 neurological patients in the Genomics England 100,000 Genome Project and identified, along with UK1-II-1 and UK2-III-2 OPDM cases, two additional individuals with an estimated repeat length of 61 and 64 repeats. We reviewed the cases and performed long read sequencing in both. The first case was a man in his 70s affected by pure hereditary spastic paraparesis (HSP), genetically unconfirmed. His brother is also affected by HSP but does not carry CCG expansion in *ABCD3*. Long read sequencing showed 120 uninterrupted CCG repeats. Upon recent evaluation he had no signs of OPDM. The second individual is a woman in her 30s affected by distal hereditary motor neuropathy and carrying a homozygous c.250G>C,p.Gly84Arg pathogenic variant in *HSPB1*. Long read sequencing of the *ABCD3* locus showed 88 CCG repeats (data not shown). In both individuals the repeat size was lower compared to the smallest pathogenic repeat identified in OPDM cases.

Finally, *ABCD3* expansion was absent from 724 control alleles and 250 alleles from internal non-OPDM samples which underwent optical genome mapping at UCL Institute of Neurology.

## SUPPLEMENTARY FIGURES

**Figure S1:**
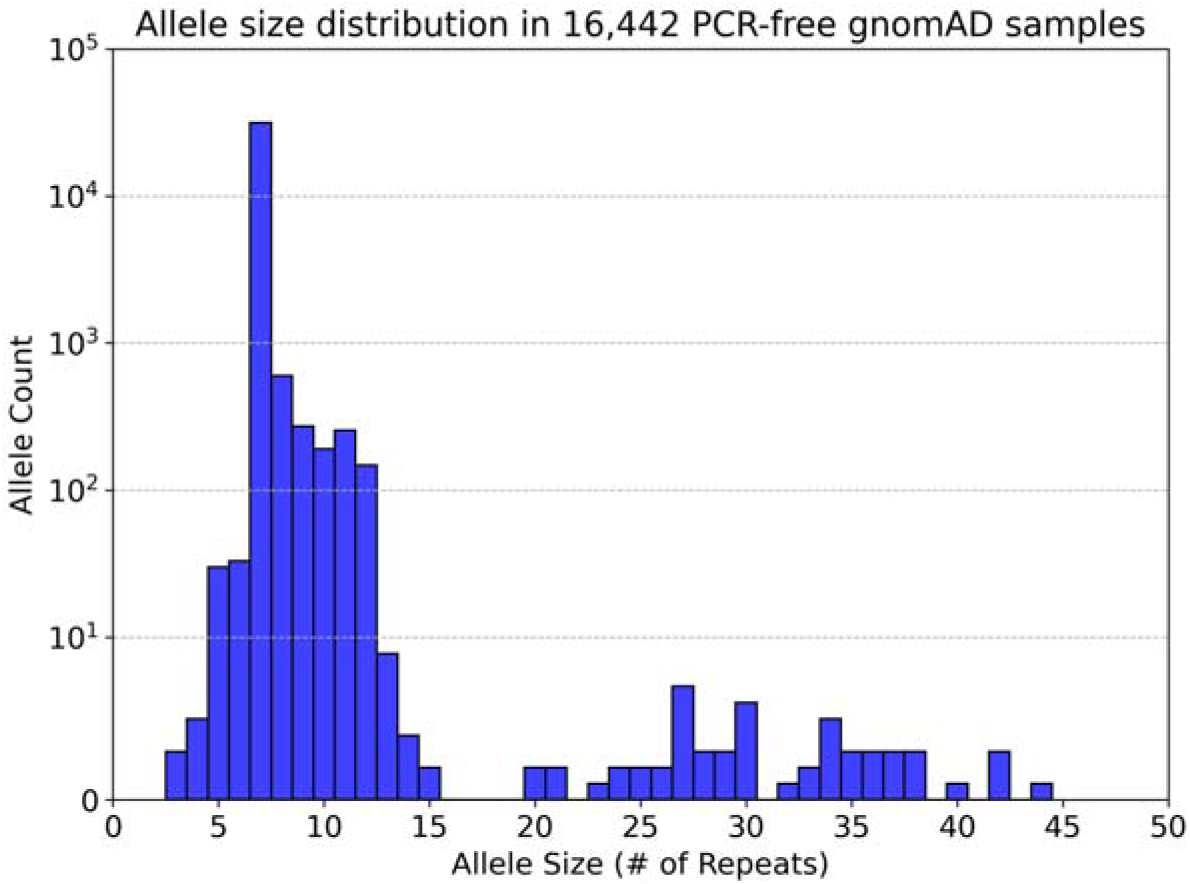
Histogram showing the distribution of *ABCD3* repeat sizes detected by ExpansionHunter in the gnomAD cohort.

**Figure S2:**
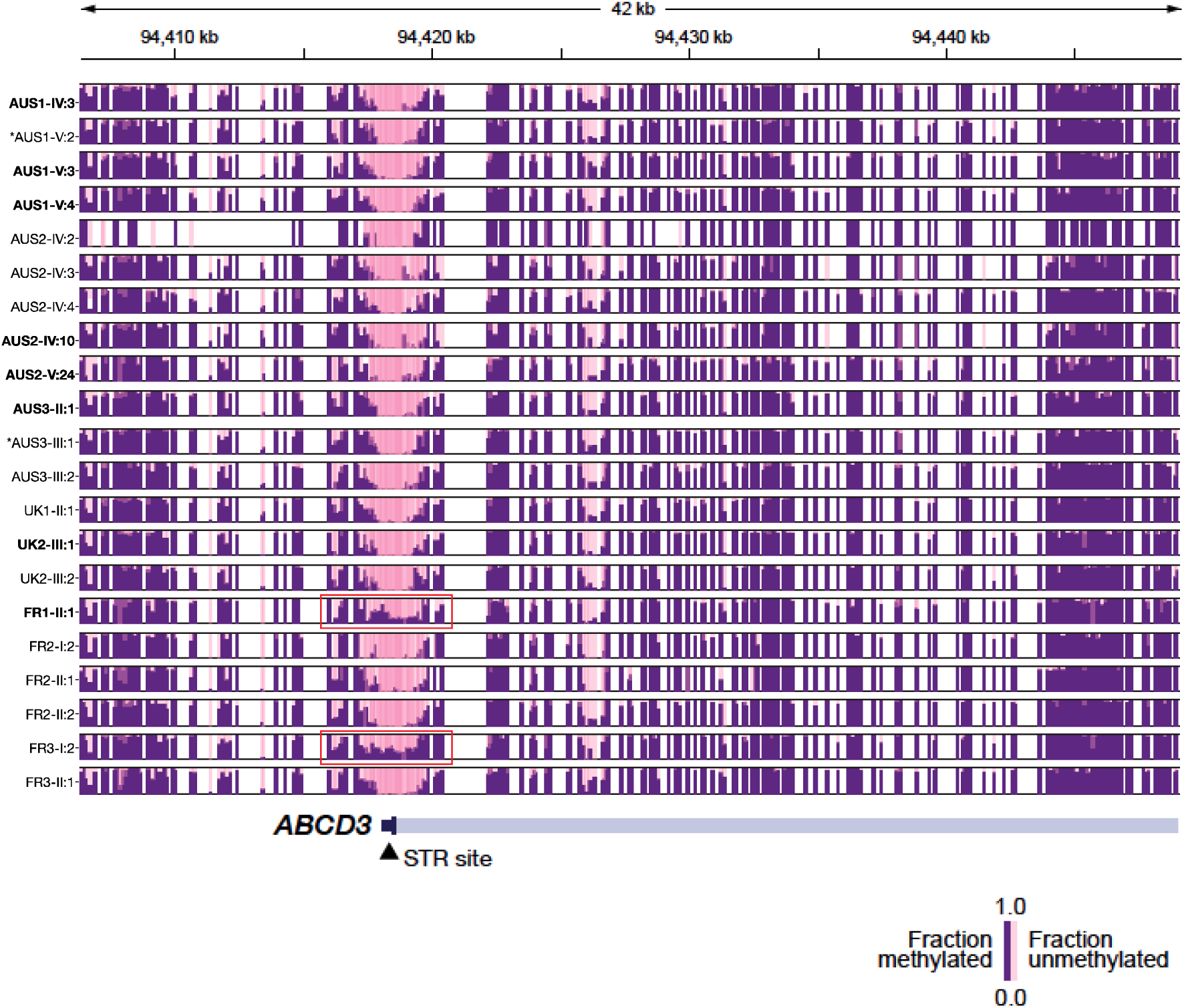
Methylation status is unchanged at the *ABCD3* promoter in 17 of 19 affected individuals with CGG expansions, compared to healthy relatives without expanded alleles (marked with an asterisk).

**Figure S3:**
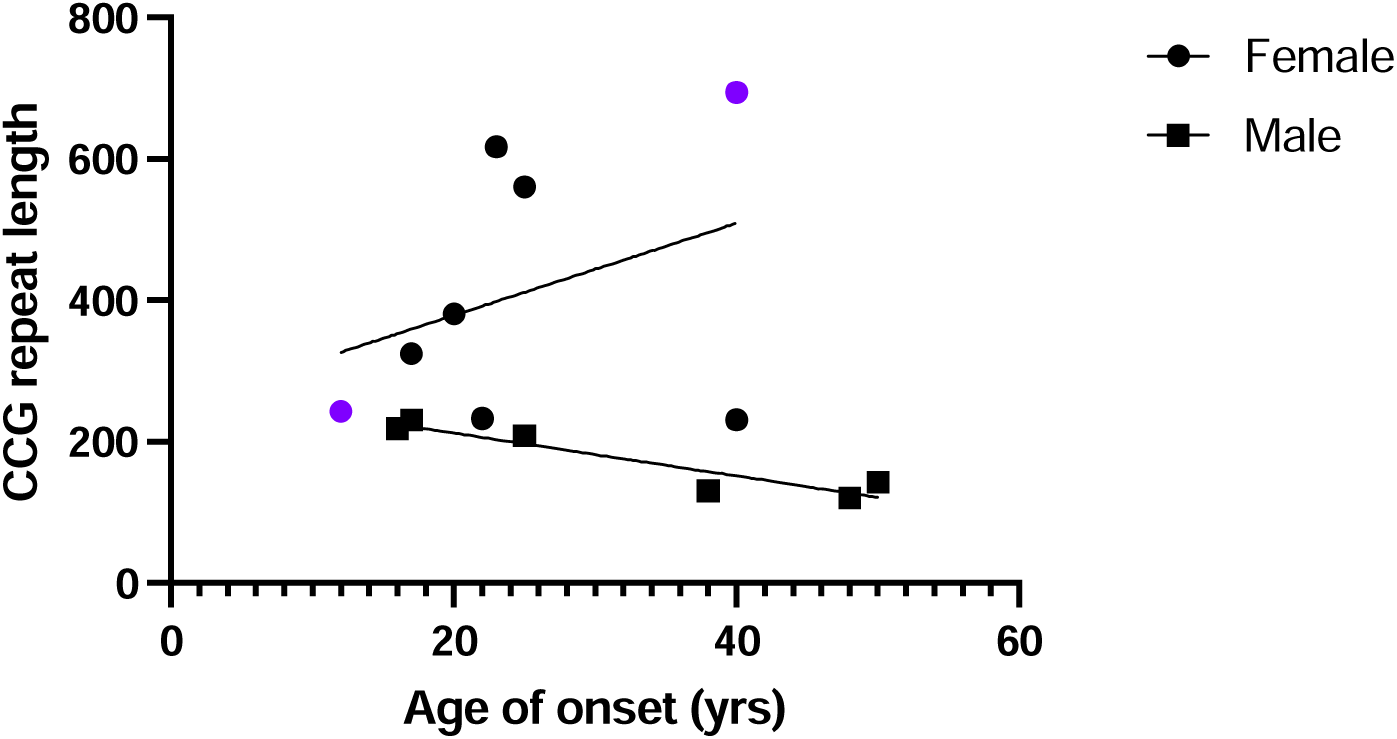
Plot of repeat size against age-of-onset in OPDM individuals, showing that larger expansions are associated with an earlier age of onset. The two females shown in purple are the individuals with hypermethylation of their expanded allele, as determined by ONT. There was a weak negative correlation between repeat expansion size and age-of-onset in affected males (*y*=3.029x+272.8, *n*=6, *p*=0.0063) with larger expansions associated with earlier onset of disease. In affected females the age-of-onset is typically ∼5 years earlier with no apparent association between repeat expansion size and age-of-onset (y=6.515x+248.3, *n*=8, *p*=0.39).

**Figure S4:**
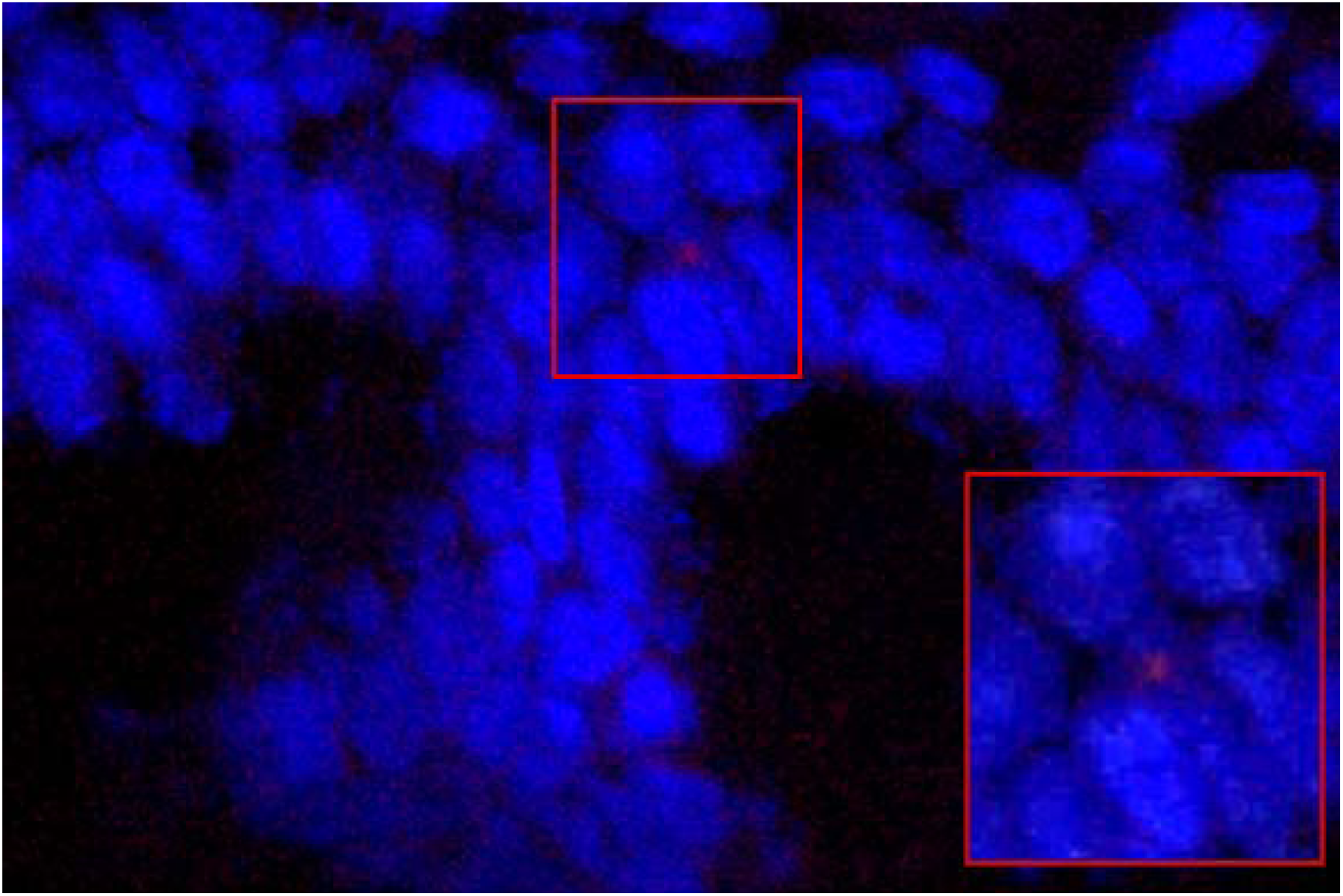
A single intranuclear CCG aggregate identified by RNA FISH in a section of fresh-frozen skin from UK2-II:1.

**Figure S5:**
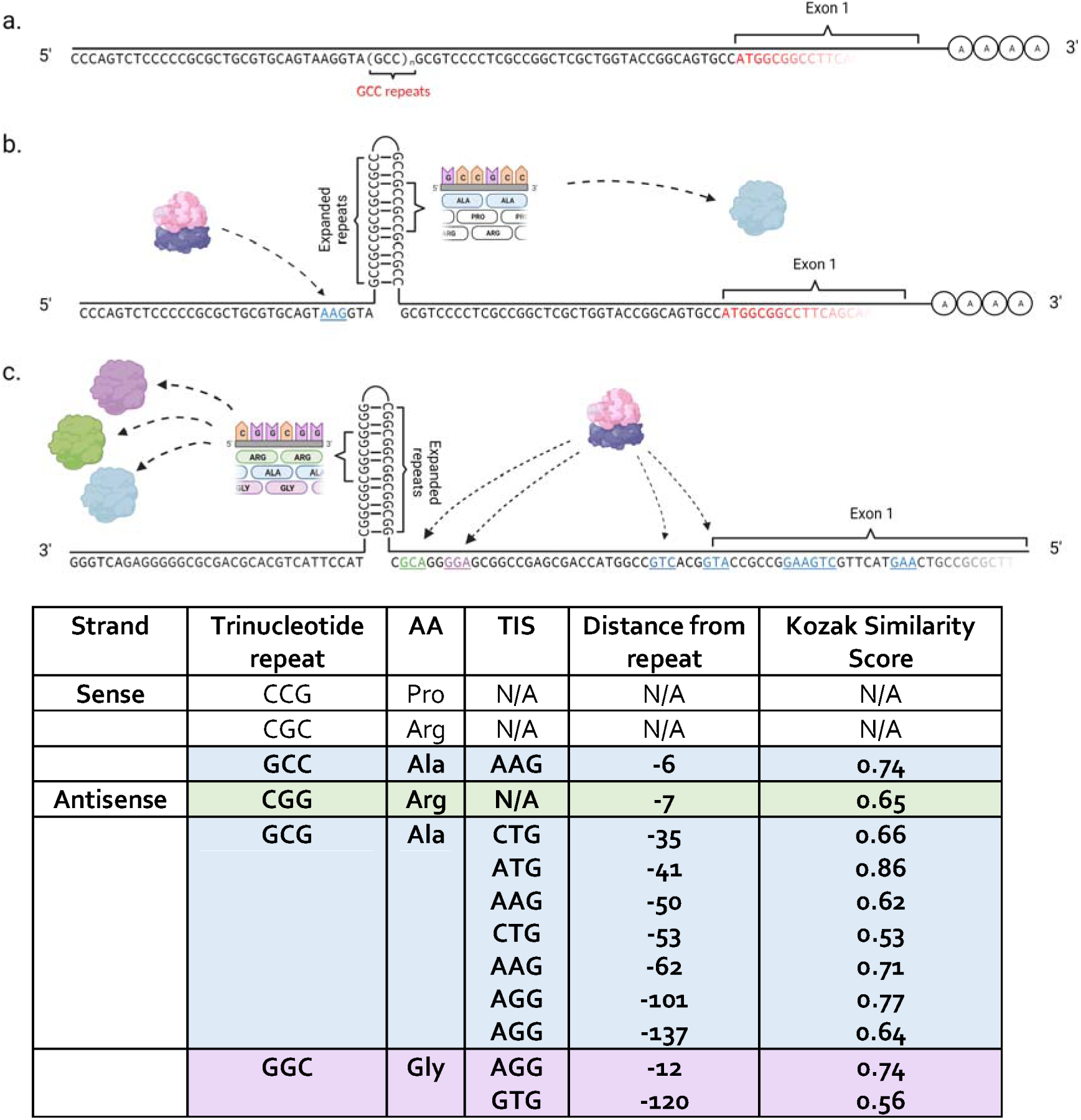
Schematic and table showing possible RAN-translation off the *ABCD3* CCG repeat expansion as predicted using the method outlined in Gleeson *et al.* 2022.^17^. 5’ sequences of *ABCD3* showing (**a**) non-expanded reference sequence mRNA with (**b**) sense strand and (**c**) anti-sense strand mRNA containing triplet repeat expansions. The translation initiation sites (TIS) are indicated for poly-alanine (light blue) and poly-glycine (green). Created with BioRender.com.

## REFERENCES

1. Satoyoshi, E. & Kinoshita, M. Oculopharyngodistal myopathy. Arch Neurol 34, 89–92 (1977).

2. Saito, R. et al. Oculopharyngodistal myopathy with coexisting histology of systemic neuronal intranuclear inclusion disease: Clinicopathologic features of an autopsied patient harboring CGG repeat expansions in LRP12. Acta Neuropathol Commun 8, 75 (2020).

3. Ogasawara, M. et al. Intranuclear inclusions in skin biopsies are not limited to neuronal intranuclear inclusion disease but can also be seen in oculopharyngodistal myopathy. Neuropathol Appl Neurobiol 48, e12787 (2022).

4. Ishiura, H. et al. Noncoding CGG repeat expansions in neuronal intranuclear inclusion disease, oculopharyngodistal myopathy and an overlapping disease. Nat Genet 51, 1222–1232 (2019).

5. Deng, J. et al. Expansion of GGC Repeat in GIPC1 Is Associated with Oculopharyngodistal Myopathy. Am J Hum Genet 106, 793–804 (2020).

6. Ogasawara, M. et al. CGG expansion in NOTCH2NLC is associated with oculopharyngodistal myopathy with neurological manifestations. Acta Neuropathol Commun 8, 204 (2020).

7. Yu, J. et al. The GGC repeat expansion in NOTCH2NLC is associated with oculopharyngodistal myopathy type 3. Brain 144, 1819–1832 (2021).

8. Yu, J. et al. The CGG repeat expansion in RILPL1 is associated with oculopharyngodistal myopathy type 4. Am J Hum Genet 109, 533–541 (2022).

9. Zeng, Y.H. et al. GGC Repeat Expansion of RILPL1 is Associated with Oculopharyngodistal Myopathy. Ann Neurol (2022).

10. Ogasawara, M. et al. Intranuclear inclusions in muscle biopsy can differentiate oculopharyngodistal myopathy and oculopharyngeal muscular dystrophy. Acta Neuropathol Commun 10, 176 (2022).

11. Yu, J., Deng, J. & Wang, Z. Oculopharyngodistal myopathy. Curr Opin Neurol 35, 637–644 (2022).

12. Beck, J. et al. Large C9orf72 hexanucleotide repeat expansions are seen in multiple neurodegenerative syndromes and are more frequent than expected in the UK population. Am J Hum Genet 92, 345–53 (2013).

13. Renton, A.E. et al. A hexanucleotide repeat expansion in C9ORF72 is the cause of chromosome 9p21-linked ALS-FTD. Neuron 72, 257–68 (2011).

14. Kovtun, I.V., Therneau, T.M. & McMurray, C.T. Gender of the embryo contributes to CAG instability in transgenic mice containing a Huntington’s disease gene. Hum Mol Genet 9, 2767–75 (2000).

15. Wheeler, V.C. et al. Factors associated with HD CAG repeat instability in Huntington disease. J Med Genet 44, 695–701 (2007).

16. Boivin, M. et al. Translation of GGC repeat expansions into a toxic polyglycine protein in NIID defines a novel class of human genetic disorders: The polyG diseases. Neuron 109, 1825–1835 e5 (2021).

17. Pettersson, O.J., Aagaard, L., Jensen, T.G. & Damgaard, C.K. Molecular mechanisms in DM1 – a focus on foci. Nucleic Acids Res 43, 2433–41 (2015).

18. Gleason, A.C., Ghadge, G., Chen, J., Sonobe, Y. & Roos, R.P. Machine learning predicts translation initiation sites in neurologic diseases with nucleotide repeat expansions. PLoS One 17, e0256411 (2022).

## SUPPLEMENTARY REFERENCES

1. Dubowitz, V., Sewry, C.A. & Oldfors, A. Muscle Biopsy – A practical approach, 4th edition, 626 (Elsevier Limited, Philadelphia, 2013).

2. Abecasis, G.R., Cherny, S.S., Cookson, W.O. & Cardon, L.R. Merlin--rapid analysis of dense genetic maps using sparse gene flow trees. Nat Genet 30, 97–101 (2002).

3. Ravenscroft, G. et al. Mutations of GPR126 Are Responsible for Severe Arthrogryposis Multiplex Congenita. Am J Hum Genet 96, 955–61 (2015).

4. Dolzhenko, E. et al. ExpansionHunter Denovo: a computational method for locating known and novel repeat expansions in short-read sequencing data. Genome Biol 21, 102 (2020).

5. Investigators, G.P.P. et al. 100,000 Genomes Pilot on Rare-Disease Diagnosis in Health Care – Preliminary Report. N Engl J Med 385, 1868–1880 (2021).

6. Payne, A. et al. Readfish enables targeted nanopore sequencing of gigabase-sized genomes. Nat Biotechnol 39, 442–450 (2021).

7. Stevanovski, I. et al. Comprehensive genetic diagnosis of tandem repeat expansion disorders with programmable targeted nanopore sequencing. Sci Adv 8, eabm5386 (2022).

8. Gamaarachchi, H. et al. Fast nanopore sequencing data analysis with SLOW5. Nat Biotechnol 40, 1026–1029 (2022).

9. Samarakoon, H. et al. Flexible and efficient handling of nanopore sequencing signal data with slow5tools. bioRxiv 10.1101/2022.06.19.496732(2022).

10. Li, H. Minimap2: pairwise alignment for nucleotide sequences. Bioinformatics 34, 3094–3100 (2018).

11. Benson, G. Tandem repeats finder: a program to analyze DNA sequences. Nucleic Acids Res 27, 573–80 (1999).

12. Gamaarachchi, H. et al. GPU accelerated adaptive banded event alignment for rapid comparative nanopore signal analysis. BMC Bioinformatics 21, 343 (2020).

13. Yepez, V.A. et al. Detection of aberrant gene expression events in RNA sequencing data. Nat Protoc 16, 1276–1296 (2021).

14. Yepez, V.A. et al. Clinical implementation of RNA sequencing for Mendelian disease diagnostics. Genome Med 14, 38 (2022).

15. Brechtmann, F. et al. OUTRIDER: A Statistical Method for Detecting Aberrantly Expressed Genes in RNA Sequencing Data. Am J Hum Genet 103, 907–917 (2018).

16. Glineburg, M.R. et al. Enhanced detection of expanded repeat mRNA foci with hybridization chain reaction. Acta Neuropathol Commun 9, 73 (2021).

17. Gleason, A.C., Ghadge, G., Chen, J., Sonobe, Y. & Roos, R.P. Machine learning predicts translation initiation sites in neurologic diseases with nucleotide repeat expansions. PLoS One 17, e0256411 (2022).

