## Supplementary material for "A CCG expansion in *ABCD3* causes oculopharyngodistal myopathy in individuals of European ancestry": Table 1

| *Table 1: Clinical findings in 24 affected individuals from eight families with a molecular diagnosis of OPDM due to CCG repeat expansions in the 5’UTR of ABCD3.* | | | | | | | | | | | | | | | | | | | |
| --- | --- | --- | --- | --- | --- | --- | --- | --- | --- | --- | --- | --- | --- | --- | --- | --- | --- | --- | --- |
| ID | Repeat length | Sex | Age at last follow up (5 y ranges) | Age of onset (5 y ranges) | First symptom | Ptosis | Ophthalmoparesis | Facial weakness | Distal LL weakness | Distal UL weakness | Proximal LL weakness | Proximal UL weakness | Dysphagia | Dysarthria | Muscle biopsy: Age (5 y range) and muscle taken | Muscle biopsy: vacuoles | Muscle biopsy: p62 intranuclear inclusions | Intranuclear inclusions on EM | CK levels (IU/L) |
| AUS1-V:3 | 130 | M | 46-50 | 36-40 | Ptosis | Yes (bilateral) | Yes | No | No | No | No | No | No | No | No | N/A | N/A | N/A | N/A |
| AUS1-V:4 | 120 | M | 46-50 | 46-50 | Ptosis | Yes (unilateral) | No | No | No | No | No | No | No | No | No | N/A | N/A | N/A | N/A |
| AUS1-IV:3 | 143 | M | 76-80 | 46-50 | Ptosis | Yes | Yes | Yes | No | No | No | No | Yes | Yes | No | N/A | N/A | N/A | N/A |
| AUS1-III:3 | N/A | F | 76-80 | 16-20 | Ptosis | Yes | Yes | Yes | Yes | Yes | Yes | Yes | Yes | Yes | 60-70, muscle  unknown | Yes | N/A | N/A | N/A |
| AUS3-II:1 | 230 | M | 66-70 | 16-20 | Ptosis | Yes | Yes | Yes | Yes | Yes | N/A | N/A | Yes | Yes | 46-50 yo, deltoid and vastus lateralis | Yes (non-rimmed vacuoles in deltoid, not in VL) | None | N/A | 321 |
| AUS3-III:2 | 129 | F | 26-30 | N/A | Ptosis | Yes | No | No | No | No | No | No | No | No | N/A |  | N/A | N/A | N/A |
| AUS2- IV:2 | 617 | F | 66-70 | 21-25 | Ptosis | Yes | Yes | Yes | Yes | Yes | Yes | No | Yes | Yes | 51-55 yo, tibialis  anterior | No | None | No | N/A |
| AUS2- V:24 | 129 | M | 21-25 | N/A | ptosis | Yes | N/A | N/A | yes | N/A | N/A | N/A | N/A | N/A | No |  | N/A | N/A | 257 |
| AUS2-IV:10 | 208 | M | 56-60 | 21-25 | Ptosis | Yes | Yes | Yes | Yes (mild) | Yes | No | No | Yes | Yes | No |  | N/A | N/A | N/A |
| AUS2-IV:3 | 324 | F | 61-65 | 16-20 | Ptosis | Yes | Yes | Yes | Yes | Yes (mild) | Yes | Yes | Yes | Yes | 56-60 yo, N/A | Yes (rimmed) | None | No | N/A |
| AUS2-IV:13 | N/A | M | 46-50 | 21-25 | Ptosis/unable to jump | Yes | Yes | Yes | Yes | Yes (mild) | Yes | No | Yes | Yes | N/A | N/A | N/A | N/A | 348 |
| AUS2-V:25 | N/A | M | 36-40 | N/A | Ptosis (mild) | Yes (mild) | No | No | No | No | No | No | No | No | No | N/A | N/A | N/A | N/A |
| AUS2-V:27 | N/A | M | 31-35 | 6-10 | Ptosis | Yes | No | Yes | Yes | Yes | No | No | Yes | N/A | No | N/A | N/A | N/A | N/A |
| AUS2-V:20 | N/A | F | 26-30 | 21-25 | Ptosis | Yes | No | No | No | Yes (mild) | No | No | No | No | No | N/A | N/A | N/A | N/A |
| AUS2-IV:14 | N/A | M | 56-60 | 36-40 | Weakness of mouth muscles | Yes | Yes | Yes | Yes | Yes | Yes (mild) | No | Yes | Yes | No | N/A | N/A | N/A | N/A |
| AUS2-IV:4 | 381 | F | 66-70 | N/A | Ptosis | Yes | N/A | Yes | Yes | N/A | N/A | N/A | N/A | N/A | 41-45 yo, deltoid | Yes | None | No | N/A |
| UK-II:1 | 381 | F | 56-60 | 16-20 | Ptosis | Yes | Yes | yes | yes | Yes | No | No | Yes | No | 41-45 yo, quadriceps | Yes | None | N/A | 142 |
| UK2-III:1 | 231 | F | 51-55 | 36-40 | Ptosis | Yes | Yes | Yes | Yes | Yes | Yes | No | Yes | No | N/A | N/A | N/A |  | N/A |
| UK2-III:2 | 218 | M | 46-50 | 16-20 | Ptosis | Yes | Yes | Yes | Yes | Yes | Yes | No | Yes | Yes | 31-35 yo, vastus  lateralis | No | None | N/A | 259 |
| FR1-II:1 | 243 | F | 46-50 | 11-15 | Ptosis | Yes (bilateral) | No | Yes | Yes (severe) | Yes (mild) | Yes | Yes | Yes | Yes | 26-30 yo, deltoid | Yes | N/A | No | 233 |
| FR2-II:2 | 560 | F | 66-70 | 21-25 | Ptosis | Yes (bilateral) | Yes | Yes | Yes (severe) | Yes | Yes | Yes | Yes | Yes | N/A, deltoid | Yes (in both) | N/A | N/A | 377 |
| FR2-I:1 | 118 | F | died age 81-85 | N/A | N/A | Yes | N/A | N/A | Yes | Yes | Yes | Yes | Yes | Yes | N/A | N/A | N/A | N/A |  |
| FR2-II:1 | 300 | M | Died age 61-65 | N/A | N/A | Yes | N/A | N/A | N/A | N/A | N/A | N/A | Yes | N/A | N/A | N/A | N/A | N/A | N/A |
| FR3-II:1 | 233 | F | 31-35 | 21-25 | Ptosis and nasal voice | Yes | Yes | Yes | Yes | Yes | No | No | Yes | Yes | 31-35 yo,  tibialis anterior | Yes | Yes | N/A | 289 |
| FR3-I:2 | 694 | F | 61-65 | 36-40 | Dysphagia | N/A | N/A | N/A | N/A | N/A | N/A | N/A | Yes | No | No | N/A | N/A | N/A | N/A |
